# Mental Health and Suicide Literacy among School Nurses in Japan: A Cross-Sectional Study

**DOI:** 10.1101/2025.09.30.25337020

**Authors:** Ayuko Yukawa, Sakurako Kusaka, Satoshi Yamaguchi, Takuya Arai, Fumika Sawamura, Fumiharu Togo, Tsukasa Sasaki

## Abstract

**Background:** School nurses (SN) are key providers of school health services and play a vital role in promoting adolescent mental health and preventing suicide. However, research into their mental health literacy (MHL) and suicide literacy remains limited.

**Methods:** A self-administered questionnaire survey was conducted with 337 SN from Japanese middle and high schools. The survey assessed SNs’ MHL and suicide literacy, including knowledge, attitudes, intended approaches, and confidence in addressing student mental health and suicide risks.

**Results:** One-third of SN incorrectly believed that they could manage psychotic symptoms by careful listening alone. Many hesitated to ask students about suicide plans, even when risk was evident. Over half lacked confidence in providing mental health education.

**Conclusion:** SNs’ MHL and suicide literacy are currently insufficient in Japan. Developing evidence-based training to improve these competencies is essential to strengthen school health services and promote better adolescent mental health and lower suicide risk.

## Introduction

The peak onset of many mental illnesses occurs during adolescence [1]. Mental illnesses in adolescents are associated with school refusal, academic decline, and social relationship problems, which can lead to impairments in future life [2,3]. Furthermore, mental illnesses are a major factor in suicide [4,5]. In the teenage years, suicide is a leading cause of death, with suicide being the first leading cause of death among teenagers in Japan. Taking appropriate action before or during the onset of mental disorders is crucial for preventing symptom exacerbation and saving lives [1]. However, adolescents find it difficult to recognize their own mental health problems [6], and as the severity of mental health problems increases, there is a tendency to avoid seeking help from others [7,8]. In severe cases such as having suicidal ideation, there is a tendency not to seek help [5]. Merely waiting for adolescents to seek help themselves is insufficient to prevent the exacerbation of mental distress or suicide during adolescence; it is necessary for adults in their surroundings to actively notice, enquire and provide support.

Schools are commonly places where adolescent students can initially receive mental health support [9,10]. School nurses (SN) play a crucial role in recognizing mental health issues among students, providing necessary support, encouraging students to seek medical help, and referring them to appropriate healthcare professionals [2,11-13]. To fulfil these roles, SN require mental health literacy (MHL) and suicide literacy. MHL refers to knowledge, attitudes, and skills regarding mental health [14]. Suicide literacy, a concept derived from MHL [15,16], refers to knowledge of risk factors for suicide and attitudes and responses to individuals with suicidal ideation, self-harm, or suicidal behaviour.

To our knowledge, four studies have examined the MHL and suicide literacy of SN [17-20]. Among these, two studies examined SNs’ knowledge of and attitudes to depressive symptoms in students [19,20]. However, these studies did not focus on assessing the level of MHL among SN. Another study explored SNs’ perceptions of training needs related to posttraumatic stress disorder, depression with suicidal thoughts, and psychosis [17]. However, the questionnaire only asked about symptoms in adults (aged 24–37 years) and did not investigate mental disorders or suicide risk factors common among adolescents. One study examined knowledge and attitudes regarding suicide risk [18], but among 105 participants, only 26 were SN, and the results specifically for SN were not reported. Previous studies have not thoroughly examined the level of MHL and suicide literacy among SN regarding mental illnesses and suicide risk among adolescent students.

Therefore, this study aims to investigate the MHL and suicide literacy of SN. The survey examines SNs’ knowledge, recognition, attitudes to depression, schizophrenia, and social anxiety disorder (SAD), as representative mental illnesses during adolescence, and also their confidence in responding to students’ mental distress. Additionally, it assesses SNs’ appropriate attitudes and understanding levels of factors contributing to suicide risk and students exhibiting suicide-related behaviours.

## Methods

### Study Design and Procedure

This study employs a cross-sectional design. In 2021, through a local education board in Japan, a notification regarding this study was sent to all public junior and senior high schools in a prefecture, Japan, reaching out to a total of 635 SN. The prefecture was selected because it includes a large population with diverse characteristics, encompassing both urban and rural areas, which provides a sample considered representative of Japan.

Of the SN contacted, 337 submitted consent forms and completed the survey, resulting in a participation rate of 53% (Figure 1). Reasons for non-participation were not collected. At the beginning of the questionnaire, the purpose and details of the study and the intended use of the data were explained in writing. School nurses were asked to indicate their willingness to participate by selecting “yes” or “no.” Those who answered “yes” proceeded to complete the survey, and this response was regarded as providing written informed consent. The study targeted professional school nurses and did not involve minors. The study protocol was approved by the Ethics Committee of the Graduate School of Education, The University of Tokyo (approval no. 18-48).

**Figure 1.**
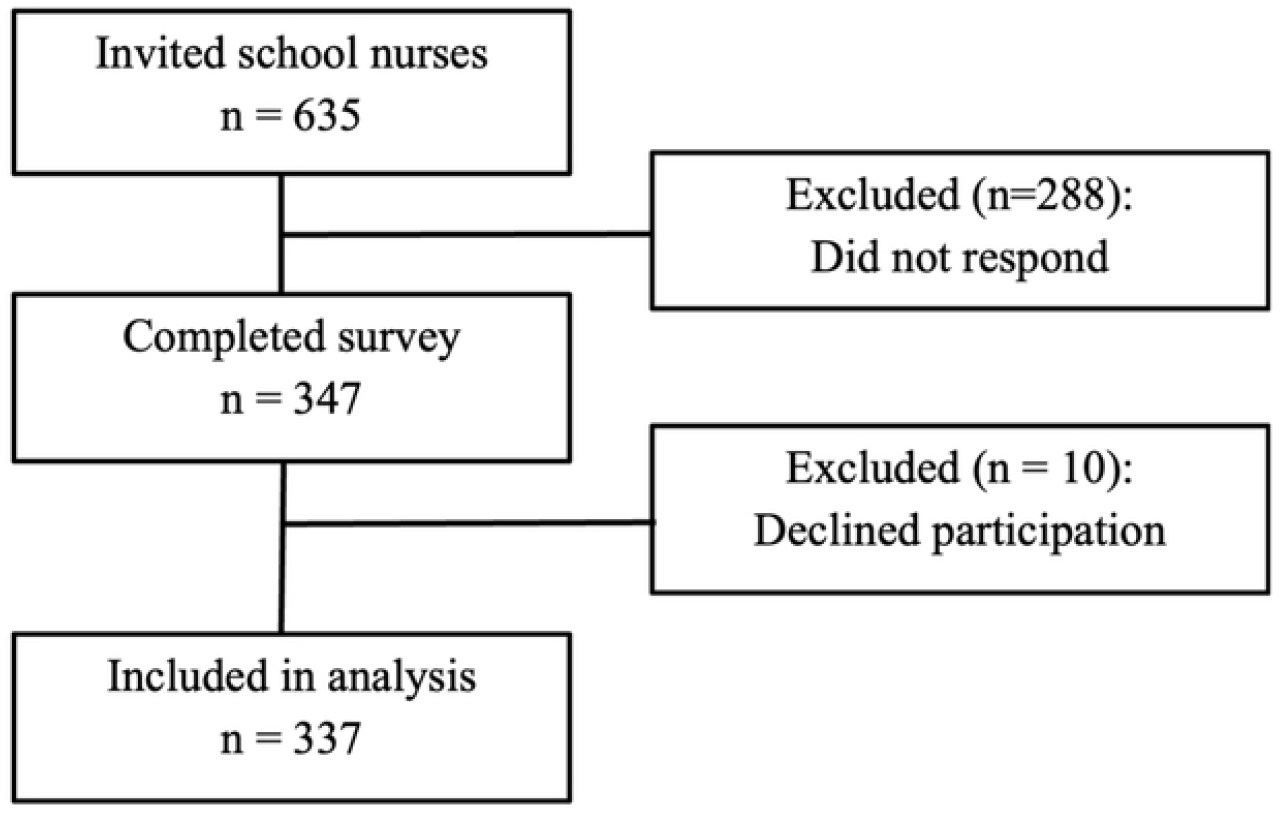
Participant flow diagram.

### Demographic Characteristics of Participants (Table 1)

Among the participants, 65% were aged 40 or above, and 54% had over 20 years of experience as SN. The majority held university degrees (74%), had experience in supporting students with mental disorders (87%), and had undergone mental health-related training in the past (79%). The sample size was based on the number of SN who voluntarily agreed to participate, and no formal sample size calculation was conducted.

**Table 1.**
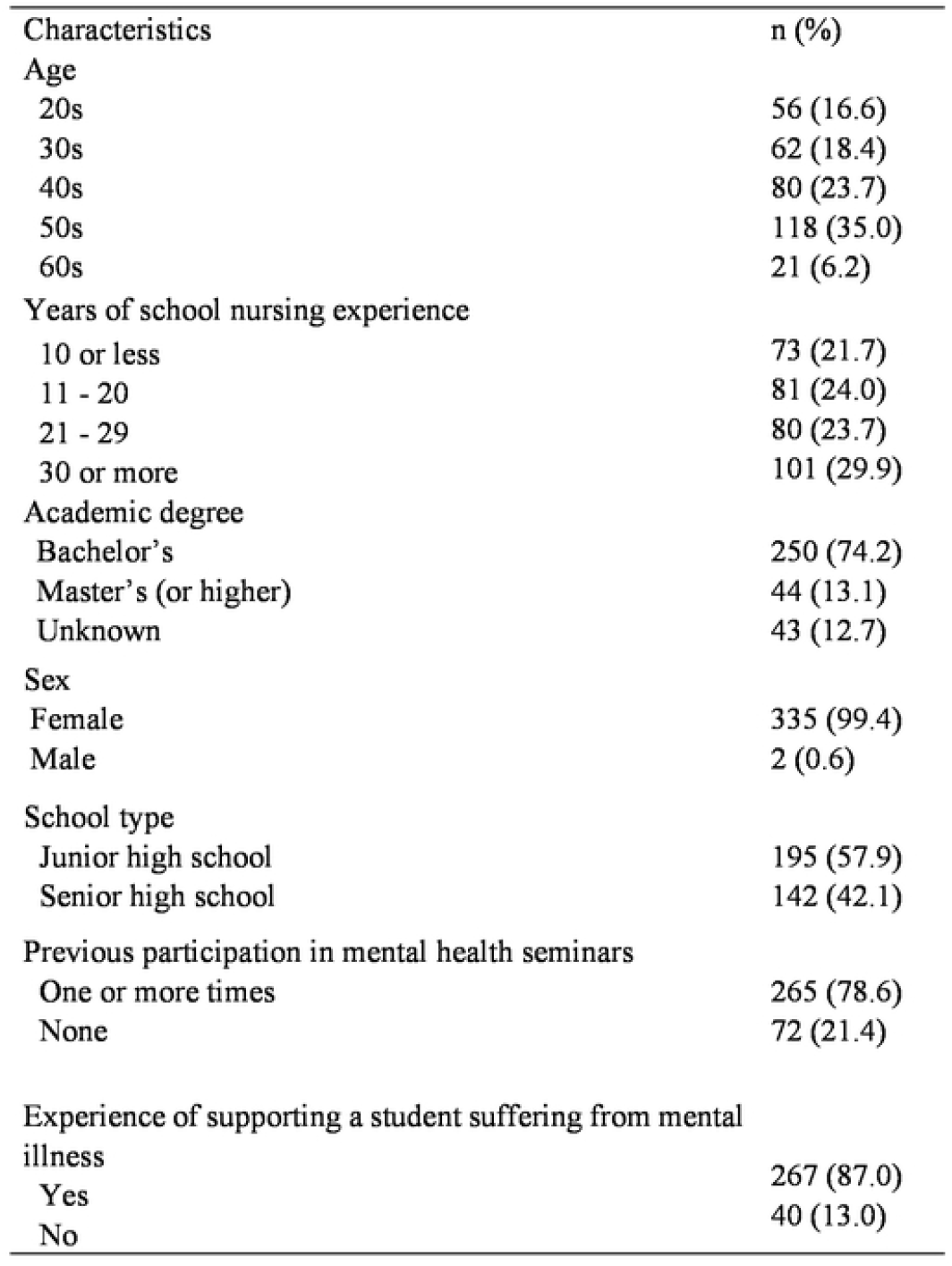
Demographic characteristics of participants.

| Characteristics | n (%) |
| --- | --- |
| Age |  |
| 20s | 56 (16.6) |
| 30s | 62 (18.4) |
| 40s | 80 (23.7) |
| 50s | 118 (35.0) |
| 60s | 21 (6.2) |
| Years of school nursing experience |  |
| 10 or less | 73 (21.7) |
| 11 - 20 | 81 (24.0) |
| 21 - 29 | 80 (23.7) |
| 30 or more | 101 (29.9) |
| Academic degree |  |
| Bachelor's | 250 (74.2) |
| Master's (or higher) | 44 (13.1) |
| Unknown | 43 (12.7) |
| Sex |  |
| Female | 335 (99.4) |
| Male | 2 (0.6) |
| School type |  |
| Junior high school | 195 (57.9) |
| Senior high school | 142 (42.1) |
| Previous participation in mental health seminars |  |
| One or more times | 265 (78.6) |
| None | 72 (21.4) |
| Experience of supporting a student suffering from mental illness |  |
| Yes | 267 (87.0) |
| No | 40 (13.0) |

### Assessment of Mental Health Literacy (MHL) and Suicide Literacy

MHL and suicide literacy were evaluated using self-administered questionnaires. The questionnaires were developed and refined by a team of psychiatrists, psychologists, educators, and school nurses. The questionnaire consisted of seven parts:

#### Part 1: Demographic Variables

This section included questions regarding age, gender, school level (junior high or high school), years of experience as a school nurse, educational background, participation in mental health-related training, and experience of supporting individuals with mental disorders (Table 1).

#### Part 2: Knowledge about Mental Health

Basic knowledge about symptoms and treatments of mental illnesses commonly observed during adolescence, such as depression, schizophrenia, and SAD, was assessed using ten questions (see Table 2). The questions were based on Japan’s demographic statistics and were developed by the authors (a team including psychiatrists, psychologists, schoolteachers and SN). Topics were chosen to determine the extent to which SN understood the importance of attention to and care and prevention of mental illness in adolescents. The internal consistency of the questions among participants in this study (Cronbach’s alpha coefficient) was 0.6.

**Table 2.** Knowledge about mental health in school nurses.

| Item | Correct answer | Proportion correct (%) |
| --- | --- | --- |
| A majority of mental disorders begin to increase in prevalence during adolescence. | T | 78.6 |
| In Japan, 1 in 5 people will suffer from some kind of mental disorder in their lifetime. | T | 50.1 |
| With mental disorders, it is undesirable to return to school before treatment is fully completed. | F | 67.7 |
| Schizophrenia develops in 1 in 100 people. | T | 52.8 |
| More than 10% of people will suffer from depression in their lifetime. | T | 59.0 |
| In some cases of mental disorders, they may only cause physical symptoms such as headaches, stomach ache or nausea. | T | 83.7 |
| By engaging in careful listening, it can be possible to cure hallucinations and persecutory delusions. | F | 71.2 |
| The duration of the treatment for depression and anxiety is usually more than a year. | T | 61.4 |
| Due to mental disorders, individuals may be unable to participate in daily conversations. | T | 92.6 |
| To reduce the risk of depression, 7 hours of sleep is best for high school students. | F | 16.3 |

Note: n = 337; T = true; F = false; Proportions indicate the percentage of correct responses for each item.

#### Part 3: Perception of Mental Health Symptoms in Vignette Cases

Participants were presented with vignettes depicting symptoms of depression, schizophrenia, and SAD commonly seen in adolescents. The vignettes were developed by the authors (the same team as in part 2). SN were asked to identify which symptoms corresponded with each disorder. The vignettes described adolescents exhibiting symptoms of the respective disorders (Table 3).

**Table 3.** School nurses’ recognition of mental health symptoms in vignette cases.

| Correctly recognized the students' symptoms as: | Proportion<br>of correct<br>responses (%) |
| --- | --- |
| Depression <sup>a)</sup> | 82.5 |
| Schizophrenia <sup>a)</sup> | 82.5 |
| SAD | 83.4 |

Note: SAD = social anxiety disorder; <sup>a)</sup> Students A to C in the vignette cases (described in Methods, part 3).

The descriptions of the vignettes were as follows.

Depression: Student A complains of “headaches”, “stomach ache” and “fatigue”, and goes to the SNs’ office. A mention, “I can’t sleep well, have no appetite, am unable to enjoy my favourite TV shows, and I can’t concentrate on my studies.” Recently, A’s tardiness has been increasing.

Schizophrenia: Student B appears to be unable to concentrate during classes. B covers her/his ears during break times. When you (SN) ask B about her/his condition, B mentions, “Everyone in the class is talking about me. Someone is always watching me. Strangers passing by say hurtful things to me. I’m extremely bothered by the situations surrounding me.”

SAD: Student C is extremely introverted. C can talk with her/his family, but cannot communicate well with anyone else, and struggles with communication with classmates. C can hardly make friends even though she/he wants to. During presentations in her/his class, C becomes nervous, blushes, and her/his voice may tremble. C is always worried that classmates or others may perceive her/his behaviour to be strange.

#### Part 4: Attitude to Students with Mental Health Problems

Using the same vignettes as in Part 3, participants were asked about their agreement or disagreement with statements reflecting attitudes to students with mental health problems (Table 4).

**Table 4.**
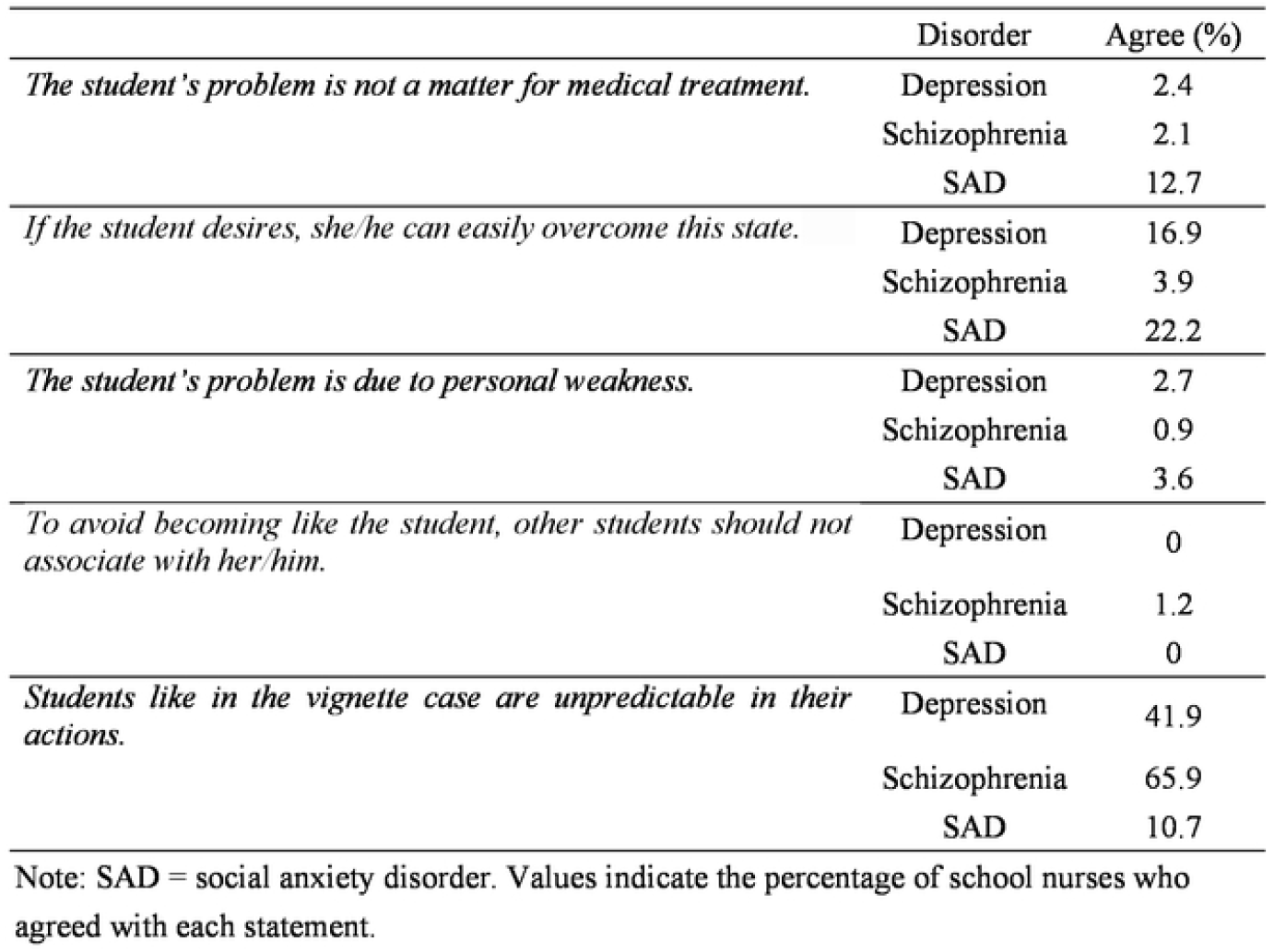
School nurses’ attitude to mental health problems in vignette cases.

|  | Disorder | Agree (%) |
| --- | --- | --- |
| *The student's problem is not a matter for medical treatment.* | Depression | 2.4 |
|  | Schizophrenia | 2.1 |
|  | SAD | 12.7 |
| *If the student desires, she/he can easily overcome this state.* | Depression | 16.9 |
|  | Schizophrenia | 3.9 |
|  | SAD | 22.2 |
| *The student's problem is due to personal weakness.* | Depression | 2.7 |
|  | Schizophrenia | 0.9 |
|  | SAD | 3.6 |
| *To avoid becoming like the student, other students should not associate with her/him.* | Depression | 0 |
|  | Schizophrenia | 1.2 |
|  | SAD | 0 |
| *Students like in the vignette case are unpredictable in their actions.* | Depression | 41.9 |
|  | Schizophrenia | 65.9 |
|  | SAD | 10.7 |

Note: SAD = social anxiety disorder. Values indicate the percentage of school nurses who agreed with each statement.

#### Part 5: Confidence in Supporting Students with Mental Health Problems

Participants were asked to rate their confidence level in providing support and assistance to the students depicted in the vignettes (Table 5).

**Table 5.** School nurses’ confidence regarding students with mental health problems.

| Case / topic | Not at all | Not much | A little | Enough |
| --- | --- | --- | --- | --- |
| Supporting student A (depression) | 0.6 | 30.3 | 63.8 | 5.3 |
| Supporting student B (schizophrenia) | 4.2 | 49.0 | 43.6 | 3.3 |
| Supporting student C (social anxiety disorder) | 0.9 | 31.2 | 63.8 | 4.2 |
| Instructing students about mental health problems | 6.5 | 55.8 | 36.5 | 1.2 |

Note: Percentages indicate responses to each question. For vignette items (students A to C), participants were asked: "How confident would you be in providing the necessary support?"

#### Part 6: Knowledge about Suicide Risks

Basic knowledge about the epidemiology, risk factors, and care/treatment of suicidal ideation/behaviour among adolescents was assessed using ten questions (Table 6). This part was also drafted in the same manner as Part 2, and the internal consistency among participants in this study (Cronbach’s alpha coefficient) was 0.55.

**Table 6.**
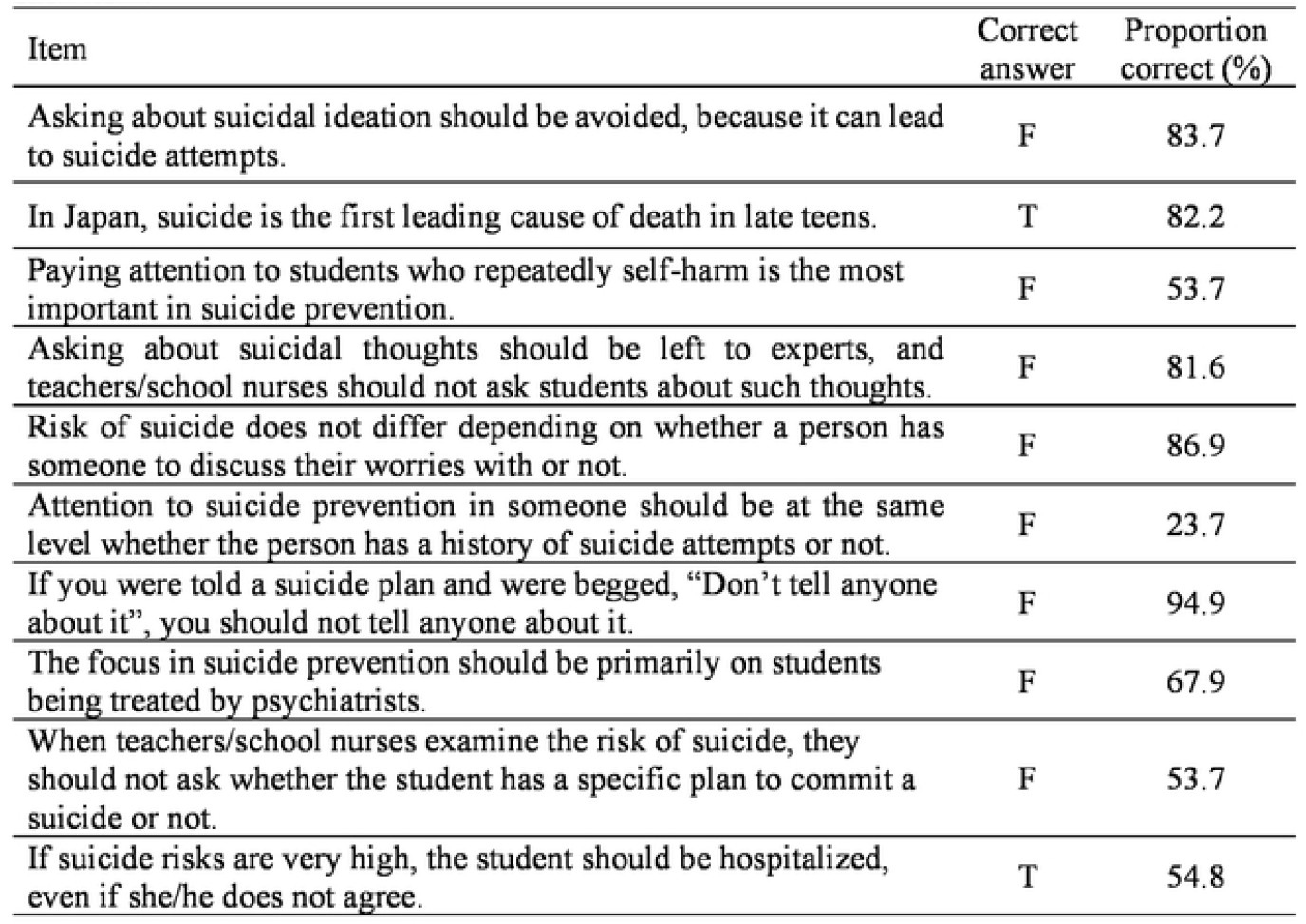
School nurses’ knowledge about preventing suicide. Note: n = 337; T = true; F = false; Proportions indicate the percentage of correct responses for each item.

| Item | Correct answer | Proportion correct (%) |
| --- | --- | --- |
| Asking about suicidal ideation should be avoided, because it can lead to suicide attempts. | F | 83.7 |
| In Japan, suicide is the first leading cause of death in late teens. | T | 82.2 |
| Paying attention to students who repeatedly self-harm is the most important in suicide prevention. | F | 53.7 |
| Asking about suicidal thoughts should be left to experts, and teachers/school nurses should not ask students about such thoughts. | F | 81.6 |
| Risk of suicide does not differ depending on whether a person has someone to discuss their worries with or not. | F | 86.9 |
| Attention to suicide prevention in someone should be at the same level whether the person has a history of suicide attempts or not. | F | 23.7 |
| If you were told a suicide plan and were begged, "Don't tell anyone about it", you should not tell anyone about it. | F | 94.9 |
| The focus in suicide prevention should be primarily on students being treated by psychiatrists. | F | 67.9 |
| When teachers/school nurses examine the risk of suicide, they should not ask whether the student has a specific plan to commit a suicide or not. | F | 53.7 |
| If suicide risks are very high, the student should be hospitalized, even if she/he does not agree. | T | 54.8 |

#### Part 7: Intended Approach to Suicidal Ideation (SI) and Suicide Plans

The content of the vignette is about a teenage student who has suicidal thoughts. The vignette modified from a previous study [21] posed the question, “If you were in a position where you interacted with the vignette student on a regular basis, would you immediately take subsequent action?” The SN were asked to respond on a four-point Likert scale of “strongly agree”, “agree”, “disagree”, “strongly disagree”. The description of the vignette was as follows.

##### Suicidal Ideation / Behaviour

“Student D feels that she/he will never be happy again and believes that her/his family would be better off without her/him. She/He has been so desperate, and she/he has been thinking of ways to end her/his life.”

Having read this vignette, SN were asked to what extent they agreed with the 5 items regarding “intention to ask students about their suicidal thoughts and plans”, and to respond on a four-point Likert scale of “strongly agree” to “strongly disagree” (see Table 7).

**Table 7.** Intention to ask about suicidal thoughts and plans.

| Item | Strongly agree | Agree | Disagree | Strongly disagree |
| --- | --- | --- | --- | --- |
| Ask whether the student does not want to live | 38.6 | 38.3 | 14.2 | 8.9 |
| Ask whether the student wants to die | 36.5 | 36.5 | 13.9 | 13.1 |
| Ask whether the student has ever thought about how to commit suicide | 32.3 | 34.4 | 21.7 | 11.6 |
| Ask whether the student has prepared for suicide | 31.5 | 35.9 | 21.4 | 11.3 |
| Ask whether the student has actually done something or been somewhere to die | 34.1 | 38.3 | 17.2 | 10.4 |

Note: n = 337. Values are percentages of responses to each item.

##### Data Analysis

Descriptive statistics including frequencies, percentages, means, and standard deviations were calculated. Multiple regression analyses were conducted to examine the relationship between mental health/suicide literacy and experience, with mental health knowledge (described in Part 2) and suicide risk knowledge (Part 6) as dependent variables and years of experience as an independent variable. Logistic regression analyses were performed to explore the relationship between confidence/intended approach and experience/literacy, with confidence in instructing students about dealing with mental health problems (Part 5) or intention to ask students about suicide (Part 7) as the dependent variables and years of experience/mental health knowledge/suicide risk knowledge as independent variables. Confidence in instructing students about dealing with mental health problems was dichotomized into “having confidence” (including answer options “A little” and “Enough”) and “lacking confidence” (including answer options “Not at all” and “Not much”). Intention to ask students about suicide was dichotomized into “intending to ask” (including answer options “Strongly agree” and “Agree”), and “Not intending to ask” (including “Strongly disagree” and “disagree”). The analysis was performed using R version 4.1.3.

Data were collected anonymously and on a voluntary basis, which may have helped mitigate potential biases such as social desirability bias and self-selection bias inherent in self-administered surveys. Cases with missing data were excluded from the analysis. Therefore, all variables included in the analyses had complete data.

## Results

### Knowledge about Mental Health (Table 2)

Approximately 1 in 5 participants did not know that “A majority of mental illnesses begin to increase in prevalence during adolescence” (79% correct). The correct response rate to the statement, “In Japan, 1 in 5 people will suffer from some kind of mental disorder in their lifetime” was 50%. Approximately 1 in 3 participants incorrectly believed that “With mental illnesses, it is undesirable to return to school before treatment is fully completed” (68% correct) and incorrectly thought that “By engaging in careful listening, it can be possible to cure hallucinations and persecutory delusions” (71% correct). Approximately 40% were unaware that “The duration of the treatment for depression and anxiety is usually more than a year” (61% correct). The item with the lowest correct response rate was “To reduce the risk of depression, 7 hours of sleep is best for high school students” (16% correct).

### Perception of Mental Health Symptoms in Vignette Cases (Table 3)

The identification rates for symptoms corresponding to depression, schizophrenia, and SAD in vignette cases depicting adolescent students suffering from mental illness were approximately 80% for each disorder. In other words, 1 in 5 participants were unable to distinguish the symptoms of mental illness.

### Attitude to Students with Mental Health Problems (Table 4)

With vignettes depicting symptoms of depression and schizophrenia, only a small percentage (2%) incorrectly agreed that these disorders are “not a matter for medical treatment”. However, with vignettes about SAD, 13% believed this not to be a matter for medical treatment. Regarding the item, “If the student desires, she/he can easily overcome this state”, 22% agreed for SAD, 17% for depression and 4% for schizophrenia. Only a small percentage of participants agreed with the statement, “The student’s problem is due to personal weakness” (3% for depression, 1% for schizophrenia, 4% for SAD). Regarding the item, “To avoid becoming like the student, other students should not associate with her/him”, 0% agreed for depression and social anxiety disorder, and 1% for schizophrenia.

### Confidence regarding Students with Mental Health Problems (Table 5)

SNs’ confidence in providing support to students exhibiting symptoms of mental illnesses (depression, schizophrenia, SAD) was as follows: 31% lacked confidence for depression, 53% for schizophrenia, and 32% for SAD. Regarding confidence in instructing students about dealing with mental health problems, 62% lacked confidence.

### Knowledge about Suicide Risks (Table 6)

Approximately half of the participants (54%) incorrectly believed that “when assessing suicide risk, it is not appropriate to enquire whether the student has a specific plan for committing suicide.” About 1 in 5 SN incorrectly believed that “Asking about suicidal thoughts should be left to experts, so teachers/school nurses should not ask students about such thoughts” (82% correct).

### Intention to Ask about Suicidal Thoughts and Plans (Table 7)

Between 1 in 5 and 1 in 3 participants expressed disagreement about asking about suicidal thoughts or plans regarding the student in the vignette case. Approximately 27% disagreed with asking whether the student wanted to die, 33% disagreed with asking whether the student had considered committing suicide, and about 28% disagreed with asking whether the student had taken steps or been somewhere with the intent to die.

### Mental Health / Suicide Literacy and Experience (Table 8)

There was no significant trend of increasing knowledge level with more years of experience. There was no statistically significant relationship between MHL and years of experience. For knowledge scores related to suicide risk, there was a trend of lower scores among SN with more years of experience, which was statistically significant (p < 0.05).

**Table 8.** Regression results for school nurses’ mental health and suicide literacy.

| Dependent variable | Independent variable | B | SE | t | p | 95% CI |
| --- | --- | --- | --- | --- | --- | --- |
| MHL <sup>a)</sup> | Experience <sup>c)</sup> | -0.0007 | 0.01 | -0.07 | p = .95 | [-0.02, 0.02] |
| Suicide literacy <sup>b)</sup> | Experience <sup>c)</sup> | -0.02 | 0.01 | -2.52 | * p < .05. | [-0.04, -0.005] |

Note: B = unstandardized regression coefficient; t = t-value; p = probability value; SE = standard error; CI = confidence interval.
<sup>a)</sup> Individual scores for Mental Health Literacy; Knowledge about Mental Health (Table 2)
<sup>b)</sup> Individual scores for Knowledge about Suicide Prevention in Adolescents (Table 6)
<sup>c)</sup> Years of school nursing experience

### Confidence / Intended Approach and Experience, Literacy (Table 9)

Regarding the item, “Confidence in instructing students about dealing with mental health problems”, responding as “confident” (choosing either “a little” or “enough”) was significantly associated with both years of experience (odds ratio (OR) 1.05) and knowledge (OR 1.32), both of which were statistically significant (p < 0.001). Additionally, the intention to enquire about suicidal ideation among students was significantly associated with both years of experience (OR 0.97) and knowledge (OR 1.51), with more years of experience correlating with lower intention (p < 0.001).

**Table 9.** Regression results for school nurses’ confidence and intended approach in relation to experience and literacy.

| Dependent variable | Independent variable | B | SE | z | p | 95% CI | OR |
| --- | --- | --- | --- | --- | --- | --- | --- |
| Confidence in instructing MHL | Experience | 0.05 | 0.01 | 3.96 | * p < .001. | [0.02, 0.07] | 1.05 |
|  | MHL | 0.28 | 0.06 | 4.48 | * p < .001. | [0.16, 0.41] | 1.32 |
| Intention to ask about suicidal thoughts | Experience | -0.03 | 0.01 | -2.61 | * p < .001. | [-0.06, -0.01] | 0.97 |
|  | Suicide literacy | 0.41 | 0.07 | 5.49 | * p < .001. | [0.27, 0.56] | 1.51 |

Note: B = unstandardized regression coefficient; z = z-value from logistic regression analysis; p = probability value; SE = standard error; CI = confidence interval; OR = odds ratio.

## Discussion

This study revealed that SN have insufficient MHL and suicide literacy, and many SN lack confidence in supporting students with mental disorders. SN with high mental health knowledge tend to have more confidence in instructing students how to deal with mental disorders, while SN with high suicide risk knowledge are more likely to have the intention to enquire about suicidal thoughts in suicidal students. These findings suggest the necessity of training SN to equip them with accurate knowledge and confidence to appropriately address the mental health and suicide risks of adolescents.

Approximately 1 in 3 SN mistakenly believed that it is undesirable for students suffering from mental illness to return to school before completing treatment, or that by engaging in careful listening, it can be possible to cure hallucinations and persecutory delusions. Additionally, about 1 in 5 SN did not know that many mental illnesses begin to increase during adolescence. This lack of knowledge can lead to the oversight of mental illnesses and create barriers to students receiving appropriate care. Furthermore, many SN believed that seven hours of sleep is sufficient for high school students, indicating that lifestyle guidance to prevent mental illness may not be appropriately provided.

It was found that many SN have insufficient knowledge and awareness regarding adolescent suicide risks. About half of the SN mistakenly believed that it is better not to ask about suicide plans, thinking it is the job of specialists, and approximately 1 in 3 SN disagreed with asking about suicidal ideation in students with suicidal thoughts/behaviour. Even if SN recognized a suicide risk, it would be difficult to prevent suicide without asking about suicidal ideation and taking appropriate measures. To prevent students from completed suicide and to provide appropriate support, it is essential to appropriately enquire about suicide risk and establish a support system tailored to the situation.

There is room for improvement in SNs’ attitudes to mental illness. 1 in 5 SN believes that individuals with SAD can easily overcome their condition if they desire to do so, while 1 in 6 holds the same belief regarding depression. Additionally, more than 1 in 10 SN do not consider SAD to be a condition that requires medical treatment. SN play a crucial role in addressing the mental health needs of students, not only within school settings but also by collaborating with community partners and general practitioners [22,23]. However, if the attitude that mental illness is not a medical issue persists, SN may be unable to fulfil their role effectively, as they may fail to refer students for medical intervention when this would be advantageous for student outcomes.

The intention to enquire about suicidal thoughts was statistically significantly related to knowledge about suicide literacy (p<0.001, OR 1.51). Confidence in instructing students about dealing with mental health problems was also statistically significantly related to the level of mental health knowledge (p<0.001, OR 1.32). These results suggest that merely accumulating experience is not enough to gain desirable knowledge and intentions; improving SNs’ knowledge and confidence through education and training is crucial.

This study highlights the need to develop training programmes that address the identified knowledge gaps: how SN should interact with students with mental disorders, the necessary medical and epidemiological knowledge, correct information about preventive lifestyles for mental disorders, and education to deepen the understanding of the importance of asking about specific suicide plans and understanding risk factors. Continuous verification and improvement of these programmes are desired to ensure their effectiveness.

## Limitations

This study targeted only SN in one prefecture in Japan. This prefecture has a large population, encompassing both urban and rural areas in the metropolitan region, and can be considered representative of Japan. However, broader regional studies with larger sample sizes are needed for generalization. The response rate was 53%, which, while not low compared with previous studies [17-20], is not high, potentially leading to participant bias.

## Conclusion

The current state in Japan of SNs’ MHL and suicide literacy is insufficient, and many SN lack confidence in dealing with students’ mental health issues. Future developments and verifications of training programmes are necessary to enable SN to confidently detect and appropriately respond to mental disorders and suicide risks early.

## Data Availability

The dataset contains potentially identifying or sensitive information of participants (school nurses). Therefore, the data cannot be made publicly available due to ethical restrictions. Data are available upon reasonable request to the corresponding author, subject to approval by the Ethics Committee of the University of Tokyo (#22-172).

## Acknowledgements

We are grateful to Ms. Michiko Nakahara from the Saitama Prefectural Education Bureau for her support in collecting data. We also extend our sincere thanks to all the school nurses who participated in the study. This work was supported by the Japan Society for the Promotion of Science (JSPS) KAKENHI Grant Number 24KJ0808.

## References

1. Solmi M, Radua J, Olivola M, Croce E, Soardo L, Salazar de Pablo G, et al. Age at onset of mental disorders worldwide: Large-scale meta-analysis of 192 epidemiological studies. Mol Psychiatry. 2022;27(1):281–95. doi:10.1038/s41380-021-01161-7

2. DeSocio J, Hootman J. Children’s mental health and school success. J Sch Nurs. 2004;20(4):189–96. doi:10.1177/10598405040200040201

3. Ligier F, Giguère CE, Notredame CE, Lesage A, Renaud J, Séguin M. Are school difficulties an early sign for mental disorder diagnosis and suicide prevention? Child Adolesc Psychiatry Ment Health. 2020;14:1. doi:10.1186/s13034-019-0308-x

4. Bridge JA, Goldstein TR, Brent DA. Adolescent suicide and suicidal behavior. J Child Psychol Psychiatry. 2006;47(3–4):372–94. doi:10.1111/j.1469-7610.2006.01615.x

5. Curtis C. Youth perceptions of suicide and help-seeking: “They’d think I was weak or ‘mental’ “. J Youth Stud. 2010;13(6):699–715. doi:10.1080/13676261003801747

6. Rickwood DJ, Deane FP, Wilson CJ. When and how do young people seek professional help for mental health problems? Med J Aust. 2007;187(S7):S35–9. doi:10.5694/j.1326-5377.2007.tb01334.x

7. Nishida A, Shimodera S, Sasaki T, Richards M, Hatch SL, Yamasaki S, et al. Risk for suicidal problems in poor-help-seeking adolescents with psychotic-like experiences: Findings from a cross-sectional survey of 16,131 adolescents. Schizophr Res. 2014;159(2–3):257–62. doi:10.1016/j.schres.2014.09.030

8. Yamaguchi S, Ando S, Miyashita M, Usami S, Yamasaki S, Endo K, et al. Longitudinal relationships between help-seeking intentions and depressive symptoms in adolescents. J Adolesc Health. 2023;73(6):1061–7. doi:10.1016/j.jadohealth.2023.06.033

9. Farmer EMZ, Burns BJ, Phillips SD, Angold A, Costello EJ. Pathways into and through mental health services for children and adolescents. Psychiatr Serv. 2003;54(1):60–6. doi:10.1176/appi.ps.54.1.60

10. Green JG, McLaughlin KA, Alegría M, Costello EJ, Gruber MJ, Hoagwood K, et al. School mental health resources and adolescent mental health service use. J Am Acad Child Adolesc Psychiatry. 2013;52(5):501–10. doi:10.1016/j.jaac.2013.03.002

11. Arnold JL, Baker C. The role of mental health nurses in supporting young people’s mental health: A review of the literature. Ment Health Rev J. 2018;23(3):197–220. doi:10.1108/MHRJ-09-2017-0039

12. Muggeo MA, Ginsburg GS. School nurse perceptions of student anxiety. J Sch Nurs. 2019;35(3):163–8. doi:10.1177/1059840517752457

13. Puskar KR, Bernardo LM. Mental health and academic achievement: Role of school nurses. J Spec Pediatr Nurs. 2007;12(4):215–23. doi:10.1111/j.1744-6155.2007.00117.x

14. Jorm AF, Korten AE, Jacomb PA, Christensen H, Rodgers B, Pollitt P. “Mental health literacy”: A survey of the public’s ability to recognise mental disorders and their beliefs about the effectiveness of treatment. Med J Aust. 1997;166(4):182–6. doi:10.5694/j.1326-5377.1997.tb140071.x

15. Calear AL, Batterham PJ, Trias A, Christensen H. The literacy of suicide scale: Development, validation, and application. Crisis. 2022;43(5):385–90. doi:10.1027/0227-5910/a000787

16. Ludwig J, Dreier M, Liebherz S, Härter M, von dem Knesebeck O. Suicide literacy and suicide stigma: Results of a population survey from Germany. J Ment Health. 2022;31(4):517–23. doi:10.1080/09638237.2021.1875421

17. Al-Yateem N. Mental health literacy of school nurses in the United Arab Emirates. Int J Ment Health Syst. 2018;12:6. doi:10.1186/s13033-018-0184-4

18. DeCou CR, Simeona C, Lyons VH, Rowhani-Rahbar A, Vavilala MS, Vercollone L, et al. Suicide prevention experiences, knowledge, and training among school-based counselors and nurses in King County, Washington—2016. Health Behav Policy Rev. 2019;6(3):232–41. doi:10.14485/HBPR.6.3.3

19. Haddad M, Tylee A. The development and first use of the QUEST measures to evaluate school nurses’ knowledge and skills for depression recognition and management. J Sch Health. 2013;83(1):36–44. doi:10.1111/j.1746-1561.2012.00745.x

20. Moen ØL, Skundberg-Kletthagen H. Public health nurses’ experience, involvement and attitude concerning mental health issues in a school setting. Nord J Nurs Res. 2017;38(2):61–7. doi:10.1177/2057158517711680

21. Jorm AF, Blewitt KA, Griffiths KM, Kitchener BA, Parslow RA. Mental health first aid responses of the public: Results from an Australian national survey. BMC Psychiatry. 2005;5:9. doi:10.1186/1471-244X-5-9

22. Bartlett H. Can school nurses identify mental health needs early and provide effective advice and support? Br J Sch Nurs. 2015;10(3):126–34. doi:10.12968/bjsn.2015.10.3.126

23. Hoskote AR, Croce E, Johnson KE. The evolution of the role of U.S. school nurses in adolescent mental health at the individual, community, and systems level: An integrative review. J Sch Nurs. 2023;39(1):51–71. doi:10.1177/10598405211068120

